# Establishing Reference Ranges for Vitamin D Levels in a Representative Population of Peshawar, Pakistan

**DOI:** 10.1101/2024.02.15.24302821

**Authors:** Usama, Zeeshan Rehman, Aktar Ali

**Affiliations:** Biochemistry Section, Institute of Chemical Sciences, University of Peshawar, Peshawar 25120, Khyber Pakhtunkhwa, Pakistan; Department of Biochemistry, Institute of Basic Medical Sciences, Khyber Medical University. Peshawar, Peshawar 25120, Khyber Pakhtunkhwa, Pakistan; Biological Screening Core, Warren Family Center for Drug Discovery, University of Notre Dame, Notre Dame, Indiana, USA

**Keywords:** 25-hydroxyvitamin D, Vitamin D deficiency, Parathyroid hormone, ELISA, BMI

## Abstract

**Background:** Vitamin D deficiency is prevalent in Pakistan, yet there is no consensus on the optimal range for 25-hydroxyvitamin D (25(OH)D) levels. Establishing reference ranges specific to the population is essential due to variations in age, sex, race, and diet. Parathyroid hormone (PTH) serves as a surrogate marker for vitamin D deficiency. This study aims to determine the reference range for plasma vitamin D in a healthy population in Peshawar, considering various demographic and biochemical factors.

**Methods:** A population-based cross-sectional study was conducted in four union councils of District Peshawar. Participants (n=200) aged 18-54 years underwent serum vitamin D and PTH analysis using ELISA kits. Data on sun exposure, anthropometric measurements, and dietary habits were collected.

**Results:** The majority of participants (92%) reported daily sun exposure of 15-30 minutes and had a mean BMI of 23.598±3.988 kg/m^2^. Only 28% of participants were vitamin D sufficient, while 72% were deficient. test results obtained for the reference values were 6.43–45.0 ng/mL for the percentile range of 2.5–97.5.

**Conclusion:** The high prevalence of vitamin D deficiency underscores the need for governmental and educational interventions to promote awareness and address this issue. Additionally, tailored nutrition plans are crucial to mitigate vitamin D deficiency in the local population.

## Introduction

Vitamin D is a prohormone primarily synthesized by the skin through ultraviolet B (UVB) radiation from sunlight, which converts 7-dehydrocholesterol to cholecalciferol or vitamin D. It is stored in adipose tissues or the liver. In the liver, it undergoes hydroxylation at the C-25 position to form 25-hydroxyvitamin D, which is then transported to the kidneys by a vitamin D-binding protein. Hydroxylation at the C-1 position produces the functioning biological form 1,25-dihydroxyvitamin D. The second hydroxylation step is regulated by calcium and phosphate concentrations via parathyroid hormone (PTH). Vitamin D can also be acquired from naturally occurring foods such as fatty fish, fish liver oil, and egg yolk. Adequate levels of vitamin D in the body are crucial for increasing the absorption of intestinal calcium and phosphate, thereby maintaining bone health [1].

Cross-sectional, ecological, laboratory, and observational evidence have identified that vitamin D reduces the risk of various cancers, cardiovascular diseases, diabetes, autoimmune and metabolic disorders, infectious diseases linked to decreased immunity, and even some neuropsychiatric disorders [2-4]. Studies have suggested that serum 25(OH)D levels ≥ 30 ng/ml may be necessary to maximize intestinal calcium absorption and prevent secondary hyperparathyroidism-induced skeletal conditions [5]. Vitamin D deficiency may lead to bone fractures in older adults because of calcium loss, which is correlated with secondary hyperparathyroidism. Numerous studies have shown that patients with chronic kidney disease (CKD) have a higher incidence of 25(OH)D deficiency, and CKD may worsen with the progression of vitamin D deficiency because of impairment in the de novo synthesis of the precursors to 25(OH)D [6].

Vitamin D deficiency ≥ 80% has been reported in apparently healthy general populations of densely populated cities in Pakistan [7, 8]. In all these studies, the reference range used as a cutoff between normal and deficient/suboptimal vitamin D status was based on international standards. There is no absolute consensus on the optimal range of 25(OH)D [9]. Because the reference range may vary with age, sex, race, and diet, efforts should be made to establish our own reference ranges by testing several healthy populations. Low levels of 25(OH)D initiate the production of parathyroid hormone (PTH) and may thus act as a biochemical marker for vitamin D insufficiency [10]. Studies attempting to establish local reference ranges for plasma micronutrients, including vitamin D, are scarce. Therefore, we conducted a population-based study to assess the reference range for plasma vitamin D levels in a representative sample of the healthy population of Peshawar, considering factors such as age, sex, race, diet, and plasma biochemical parameters. The secondary objective of this study was to estimate the prevalence of vitamin D deficiency among apparently healthy asymptomatic patients in a representative sample from Peshawar. Our findings suggest potential future policies that key stakeholders in Pakistan could initiate to reduce the level of vitamin D deficiency.

### Methodology

The study was conducted as a cross-sectional study encompassing four union councils within the district of Peshawar, selected randomly to define the reference range of vitamin D in an apparently healthy population. A total of 200 healthy adults aged between 18 and 40 years were recruited from the four union councils. The inclusion criteria consisted of adults who were deemed apparently healthy, free from osteomalacia or any bone diseases, malabsorption diseases, renal diseases, skin disorders, not regular users of calcium or vitamin D supplements, and belonging to a specific community within Peshawar.

Ethical clearance for this study was obtained from the Ethics Review Committee of Khyber Medical University, Peshawar, Pakistan. The recruitment of participants involved assistance from local residents. Before enrollment, subjects were provided with an information sheet (see Annexure I) detailing the purpose and components of the study, which was also verbally explained at the study site. Upon agreement, the subjects were requested to provide written informed consent (see Annexure II), which was documented by signature or thumbprint. Each participant was then assigned an anonymous study number.

Blood samples, along with dietary and anthropometric data, were collected from each participant. Questionnaires were administered to gather information on socioeconomic status, family size, number of siblings, and personal hygiene. A Health Check Questionnaire (see Annexure III) was used for preliminary data collection.

Collected blood samples were subsequently stored at -80°C until further investigation. Serum vitamin D levels were assessed using a 25-OH vitamin D enzyme-linked immunosorbent assay kit (Euroimmun, Luebeck, Germany). The reference range for 25-OH vitamin D was identified as 30 ng/ml, indicating normal levels, whereas vitamin D deficiency was defined as serum 25-OH vitamin D levels below 20 ng/ml and insufficiency as levels between 20 and 29.9 ng/ml. Serum PTH levels were analyzed using a PTH enzyme-linked immunosorbent assay Kit (Monobind ELISA kit, Accubind, USA).

### Statistical analysis

Data were analyzed using Minitab® version 17. Anderson Darling test of normality showed that the serum vitamin D data were normally distributed, hence parametric was examined. Data are expressed as mean and standard deviation. The association of serum vitamin D status with age, body mass index (BMI), nutritional intake, socioeconomic status, duration of sun exposure, and serum calcium, phosphate, cholesterol, and PTH levels was calculated using multiple regression analysis.

## Results

### Subjects & Socio-demographic characteristics

Total of 200 subjects participated in the study, among which 176 were males and 24 were females. The participants were ranged in age from 18-54 years with a mean 28.64±7.828 years. Majority of the participants (87.50%) were from middle income group. Participants (94%) were mostly educated among which 68% had higher education. Among total of the 200 participants 109 were students, 4 were house wives and 87 were job holders. Most of the subjects (92%) expose themselves to the sun for 15-30 minutes per day. Predominantly, participants were of good height and weight with a mean BMI of 23.598±3.988 kg/m^2^ and majority of the participants (63%) having normal BMI. Anthropometric characteristics of participants are further outlined in Table 1.

**Table 1:**
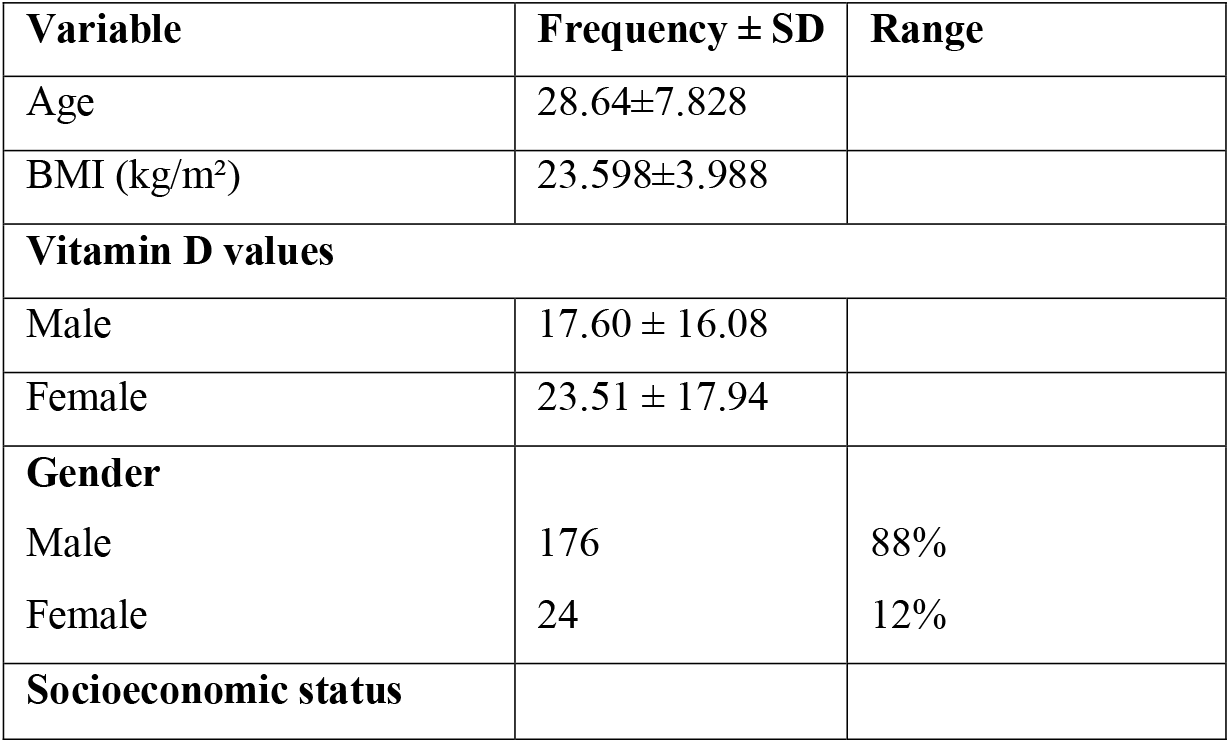
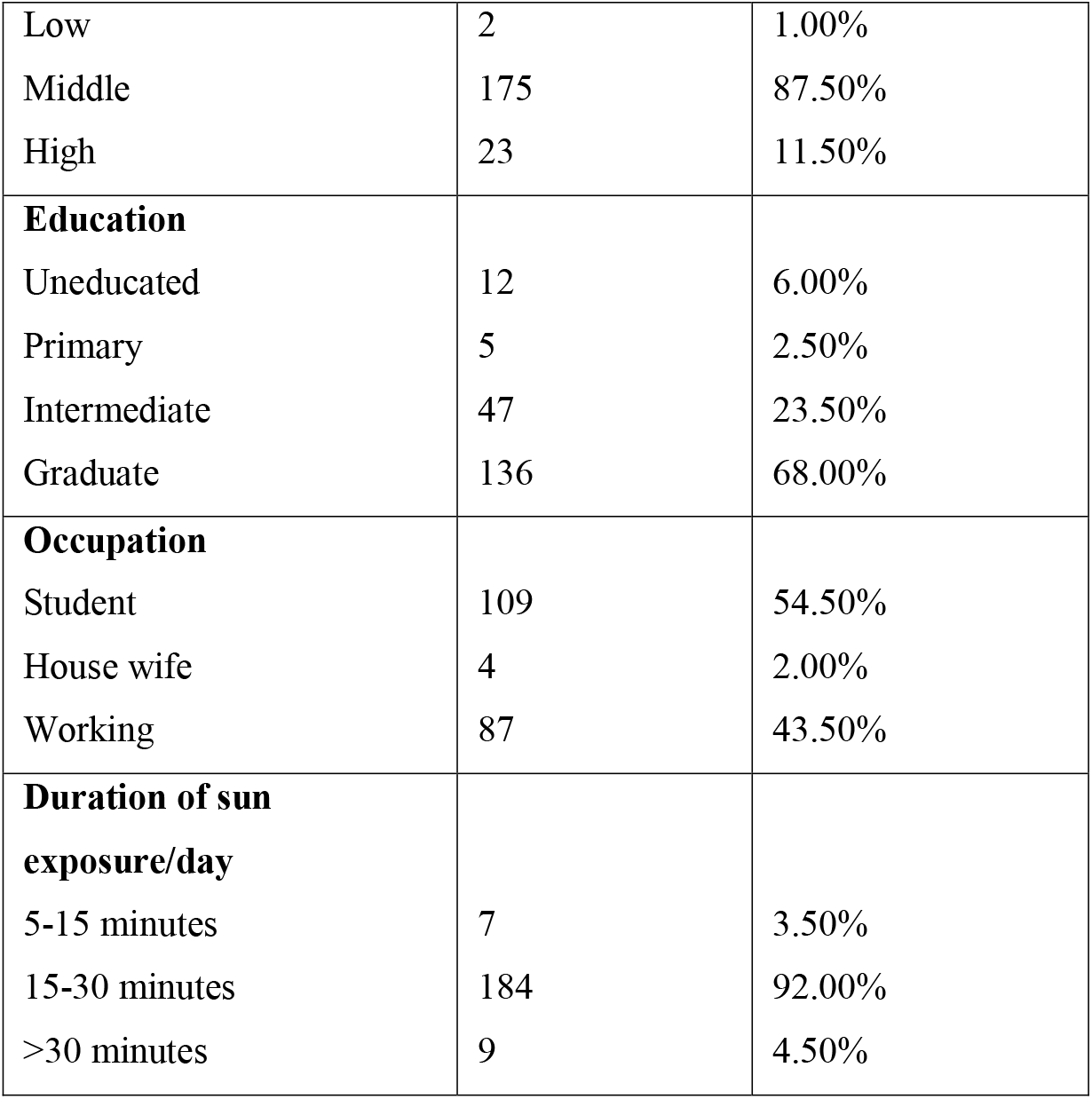
Anthropometric characteristics of study participants (n=200)

### Serum 25-OH, vitamin D status

Among total of the 200 participants, 107 (53.50%) had vitamin D deficiency (<20ng/ml), 36(18.50%) has insufficiency (>20<30) and 57 (28.50%) has normal level of serum vitamin D (Table 2).

**Table 2:** Plasma 25-OH vitamin D levels (n=200)

| **Serum vitamin D level** | **Frequency (%)** |
| --- | --- |
| Deficiency | 107 (53.50%) |
| Insufficiency | 36(18.00%) |
| Normal | 57 (28.50%) |
| Total | 200 (100.00) |

### Association of serum vitamin D with other variables

Vitamin D levels were not significantly associated with age (p=1.18), BMI (p=0.466), socioeconomic status (0.218), duration of sun exposure/day (0.786), dietary vitamin D (p=0.396), calcium (p=1.10), phosphate (p= 0.729), cholesterol (p=0.932) and PTH (0.089) levels. Total reference values are described in below table.

**Table 3:**
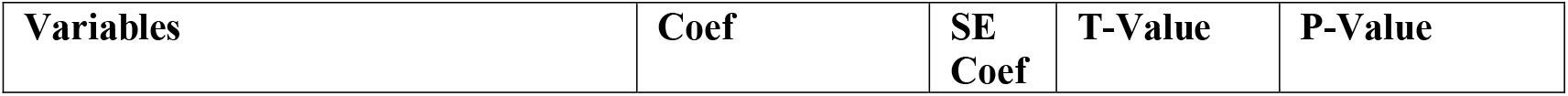
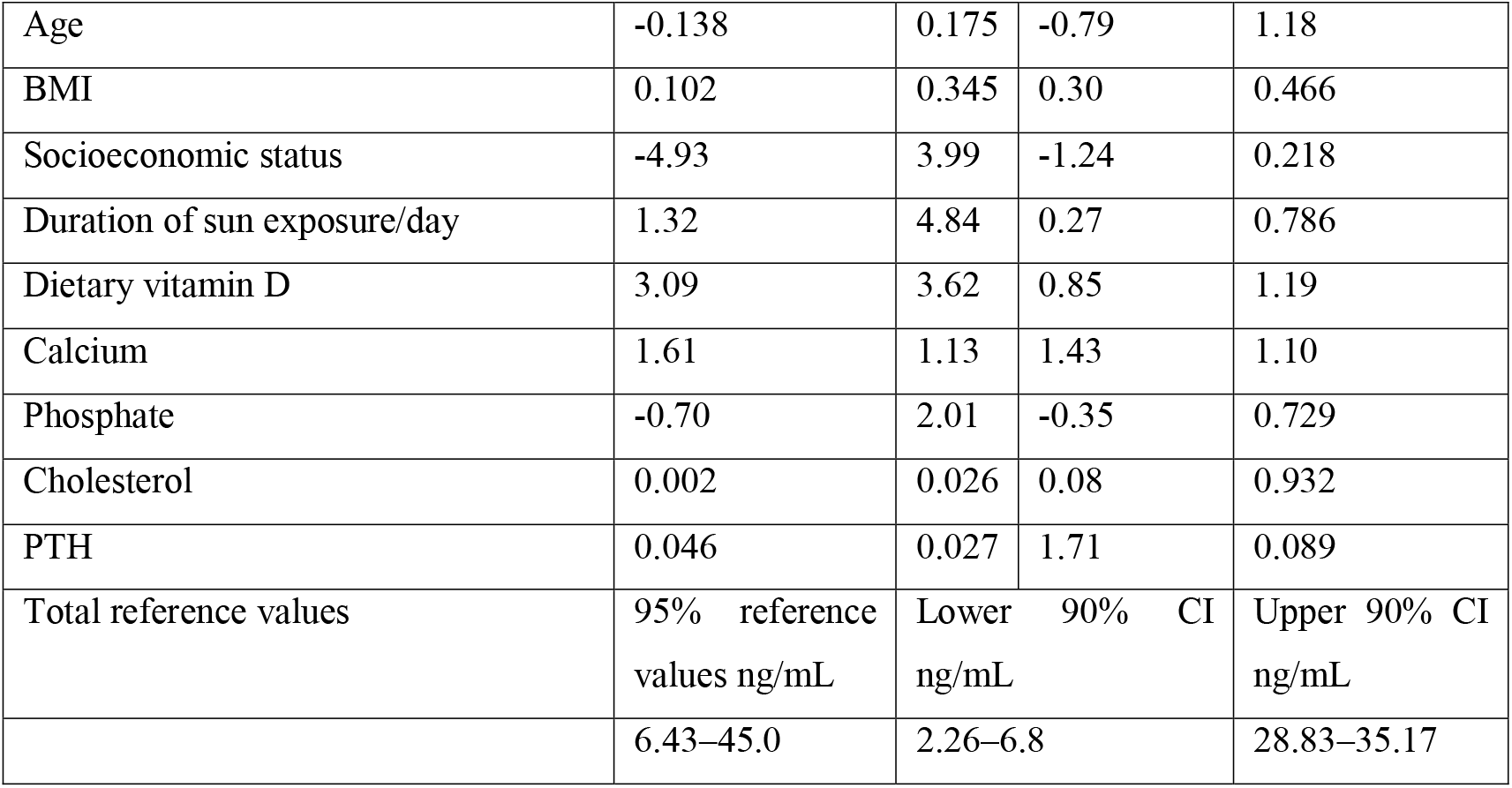
Association of serum vitamin D status with age, BMI, calcium, phosphate, cholesterol, PTH, and dietary vitamin D intake.

## Discussion

Vitamin D is a prohormone, synthesized in the skin by ultraviolet (UV) radiation of the sunlight from 7-dehdrocholesterol to cholecalciferol or vitamin D. Vitamin D can be obtained from naturally occurring food such as fatty fish, fish liver oil, and egg yolk. Some foods are however fortified with vitamin D [11]. Adequate levels of Vitamin D in the body are crucial for increasing absorption of intestinal calcium and phosphate and hence maintenance of bone health [1]. High prevalence of vitamin D deficiency (as high as 80%) has been reported in apparently healthy population of Pakistan. In all these studies, the reference range used as a cut off between normal and deficient/sub-optimal vitamin D status is based on international reference standard. There is no absolute consensus on optimal range for 25(OH) D [9]. As reference range may vary with age, sex, race, and diet; efforts should be made to establish our own reference ranges by testing a large number of healthy population [7]. Studies attempting to establish our local reference ranges for the plasma micronutrients including vitamin D are scarce. Therefore, in our study we determine the reference range for plasma vitamin D in a representative sample of the healthy population of Peshawar in relation to age, sex, race, diet and plasma biochemical parameters.

In our study almost 72% participants were either Vitamin D deficient or in-sufficient which are co-related with the previous studies conducted in Pakistan in different cities [12, 13]. Although mostly of the subjects were healthy having good BMI, and were of young age and in-addition they were mostly student/educated. Therefore, it’s obvious they will be exposed to the sunlight mostly in day-time which were also seen in the data sheet (Table 1). But due to all these variables still deficiency exists at the high rate is alarming. Our study shows no significant relation to the age, race, BMI, socioeconomic status, duration of sun exposure/day, dietary vitamin D, calcium, phosphate, cholesterol and PTH levels. In spite PTH levels are significantly corelated but the ELISA kit used for the analysis were of the poor quality and also the available machine to analysis the incubation period was also not up to the mark. May be these were the contributed reason for such a non-significant relation.

## Conclusion & Recommendations

In conclusion, reference values were 6.43–45.0 ng/mL for the percentile range of 2.5–97.5. Further, serious attention from the government and educational institutions are needed for the vitamin D sufficiency through survey or promotional ads. Also, the nutrition society needs to have, develop meals plan for the locals to overcome this major issue.

## Data Availability

All data produced in the present study are available upon reasonable request to the authors

